# Improving stroke care access through a public-private partnership, an estimation of Disability-Adjusted Life Years in patients with Ischemic Stroke undergoing mechanical thrombectomy in a IV level care center in Bogotá D.C., Colombia

**DOI:** 10.1101/2025.11.26.25341122

**Authors:** Thomas Medina, Alexandra Hoyos, Jairo Martínez, Milciades Ibáñez, Luz Adriana Camargo-López, Carlos Martínez, Jaime Rodríguez

## Abstract

**Background:** Ischemic stroke (IS) is a significant contributor to morbidity and mortality. In Colombia, there’s limited access to mechanical thrombectomy (MT). Public-private partnerships for interhospital referral may offer a solution to improve access, but their impact on health outcomes like disability-adjusted life years (DALYs) hasn’t been studied, opening possibilities to improve care and expand these strategies to other middle to low-income countries facing access barries to life-saving procedures.

**Aim:** To quantify and compare DALY accumulation from MT in patients directly treated (non-referred) at a IV level care center in Bogotá-Colombia (Fundación Santa Fe de Bogotá/FSFB) versus those referred from other centers.

**Materials and Methods:** A retrospective cohort analytical study, estimating DALY accumulation from medical records. Patients meeting inclusion criteria were included via non-probabilistic sampling. Data analysis is utilized for SPSS.

**Results:** 51 patients underwent MT, 27 were referred and 24 non-referred. The median DALY accumulation was 19.8 and 9.15 years respectively, with a significant difference between groups (p=0.003). However, after adjusting by confounding variables, including time from symptoms onset to arrival to the advance stroke center, no significant difference was found (p=0.147).

**Conclusions:** While socioeconomic and demographic differences existed between groups, adjusting by confounding variables showed comparable DALY accumulation. Prioritizing timely interhospital secondary referral strategies in acute stroke treatment is crucial, potentially offering comparable outcomes with direct care. This study provides insights into stroke management effectiveness and paves the way for future economic modeling studies justifying this care model of partnerships to guarantee access to lifesaving procedures and improve health outcomes in patients, making it a great alternative for hospitals with similar access barriers.

## Introduction

Stroke is the second leading cause of global mortality, contributing to 11.6% of total deaths and accounting for 5.7% of total DALYs worldwide (1). In Colombia, circulatory system diseases, particularly strokes, have been the leading cause of death, at 21.1% of deaths between 2005 and 2018 (2). The crude mortality rate for cerebrovascular disease in Colombia was 26.92 per 100,000 inhabitants, as the third leading cause of death (3). Stroke not only impacts mortality but also significantly contributes to disability accumulation, quality of life, productivity, and healthcare system economics (4). The economic burden of stroke is substantial, with estimated costs of $18.8 trillion globally in 2008, compounded by losses in productivity and premature mortality (5).

Socioeconomic factors are crucial in stroke prevalence, mortality, and prognosis. In Colombia, disparities in stroke prevalence and prognosis are evident across socioeconomic strata, with lower-income groups experiencing higher mortality and disability rates (6,7,8). Access to advanced therapeutic interventions like intravenous thrombolysis (IVT) or mechanical thrombectomy (MT) is limited, only a small percentage of stroke patients receive these treatments due to inadequate healthcare infrastructure and resource distribution (7,9, 10).

Timely access to MT is critical for achieving favorable stroke outcomes, demonstrated by its effectiveness within the first 12 hours of symptom onset, lack of it may lead to increased disability (11). However, the limited availability of MT-capable centers and diagnostic tools worsen delays in treatment, resulting in poorer clinical outcomes. In Colombia, most stroke patients don’t receive timely diagnosis or treatment, highlighting systemic deficiencies in stroke care delivery (10, 12).

Efficient interhospital referral systems are essential for optimizing access to endovascular reperfusion therapies for acute stroke patients (12). Collaborative networks, like the Strategic Cooperation Alliance between the Northern Subnetwork and Fundación Santa Fe de Bogotá (FSFB), a public-private partnership, facilitate timely referrals for MT-eligible patients, improving treatment coverage and clinical outcomes (13). Despite the success of such alliances, there’s a lack of studies assessing the impact of MT on DALYs lost due to stroke and its cost-effectiveness in the context of secondary referral.

This research aims to quantify and compare DALYs lost in patients treated with MT through secondary referral and those with direct care at a reference institution in Bogotá. By characterizing the study population demographically and evaluating the socioeconomic impact of MT, this study seeks to provide essential data for informing healthcare policy decision makers and expanding access to stroke treatment at a regional level.

## Methodology

### Study design

Retrospective cohort analytical design, quantitative approach and a non-probabilistic convenience sampling of patients with acute ischemic stroke (AIS) treated in FSFB. We included patients treated with MT between July 2021 and October 2022 and compared patients who consulted directly to FSFB (non-referred) with patients admitted via secondary reference (referred) from other hospitals. Patients who declined participation, failed to authorize data processing, or experienced data loss during follow-up were excluded. Ethical approval was obtained from the Corporate Research Ethics Committee at FSFB.

### Data Collection

Extensive retrieval of patient records from the hospital’s stroke center databases. Variables included in the analysis ranged from sociodemographic details (Table 1) to comprehensive clinical parameters, allowing a holistic understanding of patient characteristics and outcomes.

**Table 1.** Population description.

| Description | Total (n [%]) | Referred (n) | Non-referred (n) | P-value |
| --- | --- | --- | --- | --- |
| **Age (years)** |  |  |  |  |
| Average age | 64.47* (ST 16.67) | 56.19* (ST 14.23) | 73.79* (ST 14.28) | < 0.001' |
| Total # of patients | 51 (100%) | 27 | 24 |  |
| **Gender** |  |  |  |  |
| Female | 33 (64.7%) | 15 (55.6%) | 18 (75.0%) |  |
| Male | 18 (35.2%) | 12 (44.4%) | 6 (25%) |  |
| total | 51 (100%) | 27 | 24 | 0.147'' |
| 1 | 1 | 1 (4.2%) | 0 (0%) |  |
| 2 | 11 | 10 (41.7%) | 1 (4.3%) |  |
| 3 | 17 | 12 (50%) | 5 (21.7%) |  |
| 4 | 11 | 1 (4.2%) | 10 (43.5%) |  |
| 5 | 4 | 0 (0%) | 4 (17.4%) |  |
| 6 | 3 | 0 (0%) | 3 (6.4%) |  |
| total | 47 (92.2%) | 24 | 23 | < 0.001''' |
| Missing | 4 (7.8%) |  |  |  |
| **Highest level of education** |  |  |  |  |
| Some primary | 6 | 5 (18.5%) | 1 (4.2%) |  |
| Full primary | 10 | 7 (25.9%) | 3 (12.5%) |  |
| Some secondary | 2 | 2 (7.4%) | 0 (0%) |  |
| Full Secondary | 14 | 7 (25.9%) | 7 (29.2%) |  |
| Incomplete Vocational education | 0 | 0 (0%) | 0 (0%) |  |
| Full Vocational Education | 3 | 3 (11.1%) | 0 (0%) |  |
| Incomplete undergraduate | 1 | 0 (0%) | 1 (4.2%) |  |
| Undergraduate | 10 | 1 (3.7%) | 9 (37.5%) |  |
| Incomplete graduate level | 1 | 0 (0%) | 1 (4.2%) |  |
| Graduate level | 2 | 2 (7.4%) | 2 (8.3%) |  |
| total | 51 (100%) | 27 | 24) | 0.004''' |
| **Insurance affiliation scheme** |  |  |  |  |
| Subsidized | 12 | 10 (37%) | 2 (8.3%) |  |
| Contributory | 24 | 14 (51.9%) | 10 (41.7%) |  |
| Special | 6 | 2 (7.4%) | 4 (16.7%) |  |
| Pre-paid medicine | 6 | 0 (0%) | 6 (25%) |  |
| Direct payment by the individual | 3 | 1 (3.7%) | 2 (8.3%) |  |
| total | 51 (100%) | 27 | 24 | 0.007"" |
| **Employment status** |  |  |  |  |
| Unemployed | 6 | 4 (14.8%) | 2 (8.3%) |  |
| Retired | 8 | 3 (11.1%) | 5 (18.5%) |  |
| Self-employed | 7 | 5 (18.5%) | 2 (8.3%) |  |
| Employed | 12 | 7 (25.9%) | 5 (20.8%) |  |
| Homemaker | 18 | 8 (29.6%) | 10 (41.7%) |  |
| total | 51 (100%) | 27 | 24 | 0.622"" |
| **Monthly income (in Monthly Minimum Wages)** |  |  |  |  |
| <1 | 6 | 6 (42.9%) | 0 (0%) |  |
| Up to 1 | 3 | 3 (21.4%) | 0 (0%) |  |
| 1 – 2 | 4 | 2 (14.3%) | 2 (25%) |  |
| More than 2 | 9 | 3 (21.4%) | 6 (75%) |  |
| total | 22 (43.1%) | 14 | 8 | 0.27"" |
| Missing | 29 (56.9%) |  |  |  |
| **Housing** |  |  |  |  |
| Homeownership | 16 | 9 (64.3%) | 7 (87.5%) |  |
| Rental | 4 | 4 (28.6%) | 0 (0%) |  |
| Another | 2 | 1 (7.1%) | 1 (12.5%) |  |
| total | 22 (43.1%) | 14 | 8 | 0.342"" |
| Missing | 29 (56.9%) |  |  |  |

\* = Mean. SD = Standard Deviation. ' = Two-tailed t-test. " = Asymptotic Pearson chi-squared test (two-tailed). "" = Exact Mann-Whitney test (two-tailed). "" = Exact Pearson chi-squared test (two-tailed). "" = Exact Fisher test.

### Statistical Analysis

Descriptive statistics were used to characterize the final sample, offering insights into the distribution of key variables. Parametric tests, like the independent two-group Student’s t-test, were applied to normally distributed data, while non-parametric tests, including the Wilcoxon-Mann Whitney test, were utilized for variables deviating from normality. Multivariate analysis, facilitated by ordinal non-parametric regression, explored the influence of various confounding factors on the outcomes of interest. Statistical significance was set at p < 0.05, with a confidence interval of 95%. SPSS 25.0 software was used to perform statistical analysis.

### Bias Control

Expert selection of MT candidates and inter-hospital referrals mitigated inherent selection bias. Acknowledging the study’s single-center focus, efforts were made to ensure internal validity by comparing DALY accumulation and cost-effectiveness ratio using real billing data. Bivariate and multivariate analyses considered various influencing variables, although limited sample size and incomplete data accessibility posed challenges.

Challenges were encountered in accurately determining hospitalization costs and capturing complete complication details, attributed to restricted access to information and patients completing their stays at other facilities. Cost data extracted from direct billing rather than micro-costing introduced biases, yet efforts were made to address these during result analysis to draw meaningful conclusions.

## Results

### Demographic Characteristics

The study included 51 patients who underwent MT at a level IV care center. 27 patients (52.9%) were the referred group, 24 patients (47.1%) were the non-referred group treated directly at FSFB. The average age was 64.47 years. The referred group had an average age of 56.19 years, significantly lower than the non-referred group’s average age of 73.79 years (p < 0.001). There were no significant gender differences between the groups (p= 0.147). 70% of the referred patients came from the Northern Health Services Subnetwork.

Significant differences were found between the referred and non-referred groups in terms of socioeconomic (p < 0.001) and educational level (p=0.004), as well as affiliation regime to the health system (p=0.007). However, there were no statistically significant differences in employment status, monthly income level, or type of housing between the groups, albeit with limited access to information for the latter two variables (data from 43% of the sample) (Table 1).

### Clinical Characteristics

The non-referred group had a higher prevalence of hypertension (p= 0.035), and DM2 (p= 0.016) compared to the referred group. Conversely, alcohol consumption was more common in the referred group (p=0.05). There were no significant differences in the history of obesity, smoking, dyslipidemia, atrial fibrillation, or previous stroke between the two groups (Figure 1). The baseline modified Rankin scale (mRS) was higher in the non-referred group (p=0.015),

**Figure 1.**
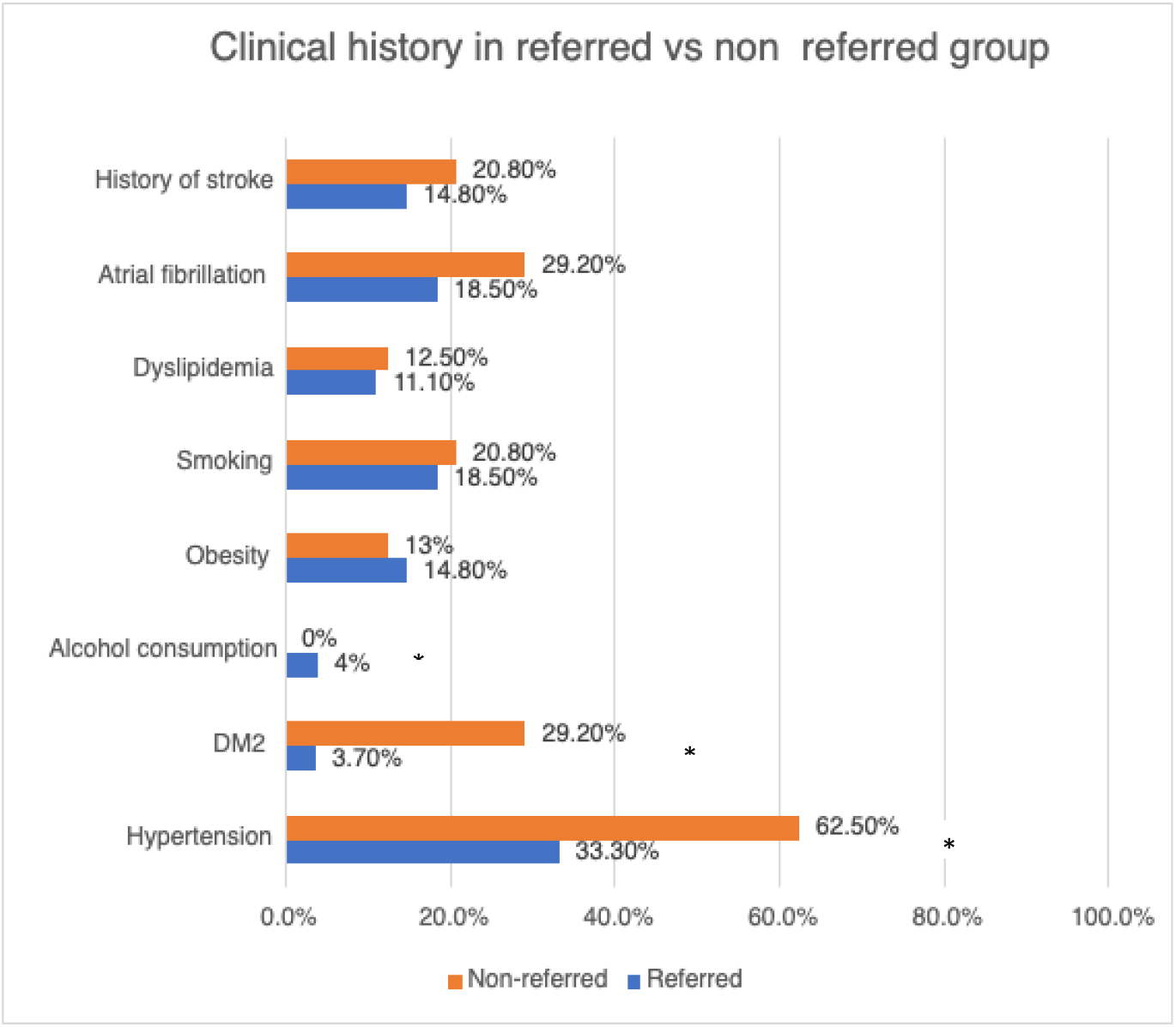
Medical history comparison between referred and non-referred groups. * = p<0.05

There were no significant differences at the end of hospitalization or in the average NIH (National Institute of Health) Stroke Scale/Score (NIHSS) NIHSS score upon admission between the groups. However, the NIHSS score upon release was higher in the referred group (p < 0.005), and the Alberta Stroke Program Early CT Score (ASPECTS) score was higher in the non-referred group (p= 0.044). Most patients presented obstruction in the left M1-M2 part of the middle cerebral artery, with no significant differences between the groups.

Regarding attention time, from *symptom onset* and *last seen well* to admission to the advanced stroke center was longer in the referred group with a statistically significant difference. While *door-to-groin* time was significantly shorter in the referred patient group, there was no significant difference in *door-to-reperfusion* time between both groups. As expected, the total time from symptom onset to reperfusion was also statistically longer in the referred patient group compared to non-referred patients (p = 0.045). In 14 of the referred patients (51.85%), it was also possible to obtain the time from IVT initiation (in the primary stroke center) to the arrival to FSFB (median time of 207 minutes), despite being relatively close with a median distance between centers of 7.5 km (SD 129.4) (Figure 2, appendix 2).

**Figure 2.**
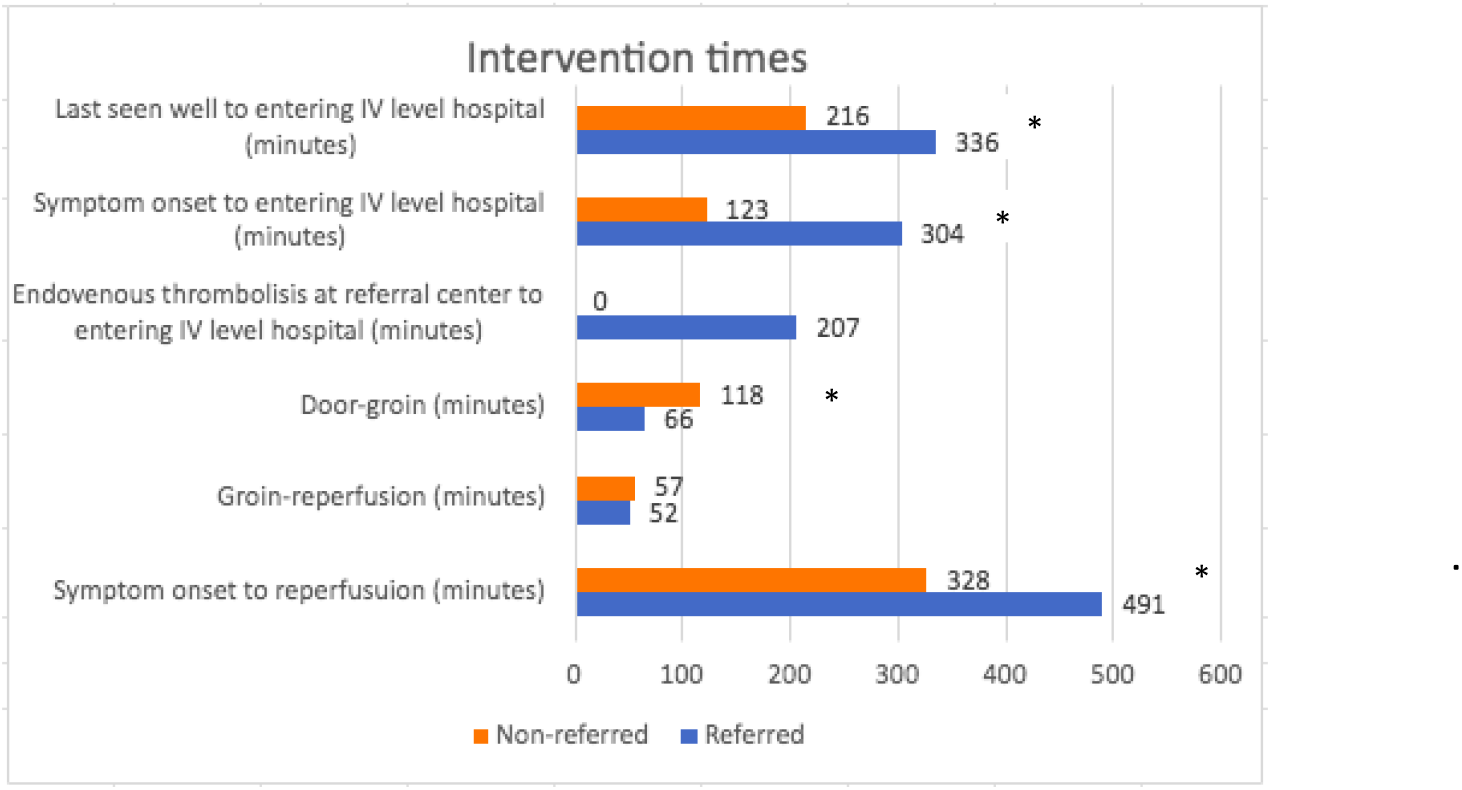
Average times of intervention between referred and non-referred groups (minutes). * = p<0.05.

There were no significant differences in successful reperfusion rate (TICI score) after MT, hospital stay duration, intensive care unit (ICU) admission rates, complications rate (during the procedure or inpatient care) nor mortality rates (Appendix 1). At the end of hospitalization, the NIHSS score was higher in the referred group (p < 0.005), and mRS was no different between the two groups.

### Cumulative DALYs and DALY-associated Cost-Effectiveness Ratio

After a median follow-up of 7 months, the mRS (52.9%), and mortality rates (62.5%) were higher in the referred group (p= 0.055 and 0.062). The cause of death was mostly attributed to ischemic stroke in both groups; systemic infections (7.1%), and other causes (28.6%) were also noted in the non-referred group (Appendix 1).

Moreover, the total accumulation of DALY lost by the entire population was 834.23 years with a mean of 16.35 years per individual. The average amount of individual DALY was higher in the “referred” group (20.833 +/-11.33 years, median of 19.801 years) compared to the “non-referred” group (11.321 +/-X6.511 years, median of 9.150 years), with a statistically significant difference (p=0.003).

Direct billing costs of care at the level IV hospital were similar between both groups, implying a higher DALY’s cost-effectiveness ratio in the non-referred group (p=0.001). Nevertheless, after adjusting for potential confounding variables, there were no statistically significant differences between DALY accumulation in the two groups (p = 0.147) (Table 2) (appendix 3). Furthermore, time from symptom onset to IV hospital admission was a crucial factor, particularly within the first 6 hours as is the standard for optimal treatment, f or a comparable outcome in terms of individual DALY burden according to the multivariable model (figure 3) (15).

**Table 2.** Disability-Adjusted Life Years (DALYs) and Costs.

| Category | Description | Total (n (%)) | Referred (n) | Non-referred (n) | P-value | Adjusted P-value |
| --- | --- | --- | --- | --- | --- | --- |
| Individual DALYs |  | 16.35* (13.89**/ SD 10.45) | 20.83* (19.80**/ SD 11.33) | 11.32* (9.15**/ SD 6.51) |  |  |
|  | total | 51 (100%) | 27 | 24 | 0.003 | 0.147+ |
| Direct billing cost (USD) |  | 20728.34* (18796.22**/ SD 9231.88) | 20783.71* (17753.24 **/ SD 11271.35) | 20666.04 * (19542.64 **/ SD 6454.12) |  |  |
|  | total | 51 (100%) | 27 | 24 | 0.125'''' |  |
| DALYs<br>Adjusted<br>Cost-Effectiveness<br>Ratio |  | 1949.21*<br>(1343.59 **/ SD<br>1633.88) | 1553.79*(896.9<br>2**/ SD<br>1775.52) | 2394.05*<br>(2155.38 **/ SD<br>1360.03) |  |  |
|  | total | 51 (100%) | 27 | 24 | 0.001'''' |  |

\*= Mean, SD = Standard Deviation, RP = Range mean. \*\* = Median. ' = Two-tailed t-test. " = Asymptotic Pearson chi-squared test (two-tailed). ''' = Exact Mann-Whitney test (two-tailed). '''' = Exact Pearson chi-squared test (two-tailed). ''''' = Exact Mann-Whitney test (one-tailed). ''''' = Asymptotic Mann-Whitney test (two-tailed). + (non-parametric ordinal regression model). ++ (non-parametric ordinal regression model) adjusted for baseline mRS, hypertension, DM2, time from symptom onset to IV level hospital triage admission, time from symptom onset to entering IV level hospital /less than 6 hours.

**Figure 3.**
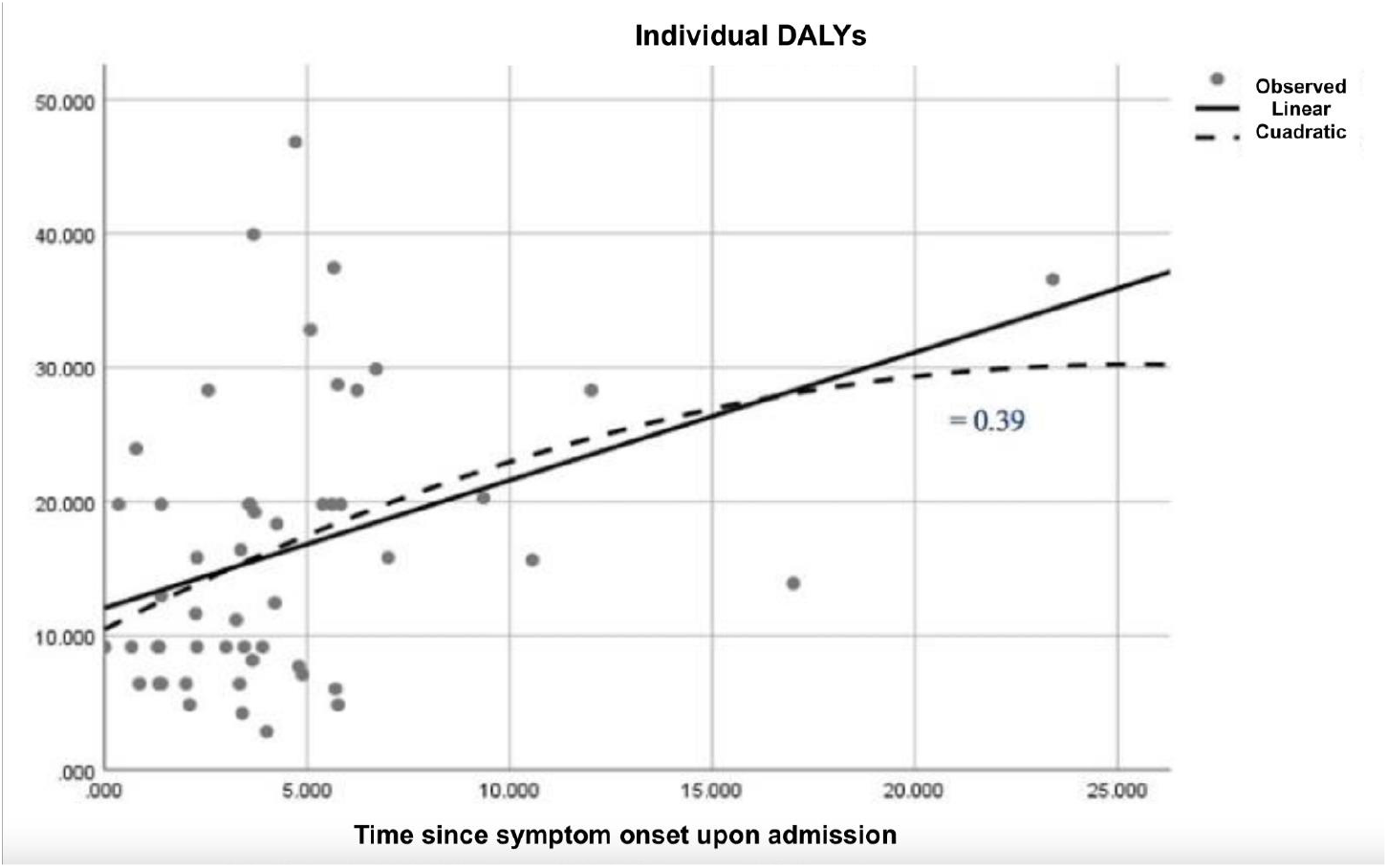
Spearman correlation model between time since symptom onset upon admission and individual average cumulative DALY burden.

## Discussion

This study assessed the cumulative DALYs burden among AIS patients (referred or non-referred) undergoing MT at a level IV facility in Bogotá D.C-Colombia. The analysis revealed a higher median DALYs burden in the referred group compared to non-referred.

Despite achieving homogeneous distribution in many clinical characteristics, the populations studied remained heterogeneous, reflecting diverse demographic, clinical, and social contexts. This heterogeneity underscores the complexity of stroke management and emphasizes the need for nuanced approaches tailored to individual patient profiles and social circumstances.

The non-referred group was notably older, leading to a higher prevalence of hypertension and DM2 within this cohort. Socioeconomic status emerged as a significant differentiator between the groups, with the non-referred patients comprising a higher proportion of contributory regime beneficiaries, high socioeconomic strata, and higher educational levels compared to the referred group.

Analysis of clinical outcomes post-MT revealed significant differences only in NIHSS scale scores at the end of hospitalization, with higher scores seen in the referred group. This discrepancy could be attributed to the longer symptom onset-to-reperfusion time and lower ASPECTS scores noted in this group. However, no differences were found in other key metrics such as post-MT TICI scores, modified Rankin scale (mRS) scores upon discharge, length of hospital stay, ICU admission rates, mortality rates, or post-MT complications. These findings suggest that MT is equally safe and effective under both direct care and inter-hospital referral contexts, but is highly dependent on times of attention, highlighting the reproducibility and feasibility of the procedure in our setting. Notably, the “door-to-groin” time was significantly shorter in the referred group, indicating efficient pre-notification and intrahospital flow processes. Likewise, the effectiveness and safety profile of MT between both care schemes reported in this study are like those reported in the literature (16,17).

The socioeconomic disparities likely played a role in influencing the long-term clinical outcomes seen in this study. Specifically, the referred group, possibly due to greater social vulnerability, may experience earlier incidence and a heavier burden of cerebrovascular disease, and lower chances of rehabilitation after the stroke, meaning worse mRS results. The worse mRS added to a younger age in the referred group probably explains the differences in DALYs. These findings align with existing literature, underscoring the association between socioeconomic factors and stroke prevalence and prognosis in Colombia. Factors such as income, education, health system affiliation, and unmet basic needs have been previously identified as determinants of stroke outcomes (6,7,8).

This study’s main contribution lies in the identification of a higher average cumulative DALY burden in the referred group compared to the non-referred group. This difference in burden significantly affected the adjusted cost-effectiveness ratio, favoring direct care primarily due to DALY burden asymmetry. Further analysis revealed that the time from symptom onset to IV level hospital admission was a crucial factor, particularly within the first 6 hours, which is considered an optimal timeframe for intervention initiation (15) (figure 3). These findings align with existing literature, which emphasizes the importance of prompt intervention and optimized referral processes in stroke care. Specifically, studies that compared the non-referred vs referred model of stroke care have shown comparable results with shorter symptom onset-to-reperfusion times in the referred model (16), underscoring the critical role of efficient referral processes between hospitals to stroke management.

Despite the potential benefits of inter-hospital referral, current practices in our region are hindered by administrative and logistical delays. Addressing these challenges is essential for improving stroke care and improving patient outcomes. While this study offers valuable insights into stroke management strategies in our setting, it is not without limitations. The heterogeneity of the two groups hinders the ability to compare the effects of the two modalities of admission on DALYs. Additionally, the small sample size limits the generalizability of our findings, and the single-center nature of the data exposes the study to potential selection bias. Lastly, incomplete data for certain variables and potential recall bias among surveyed patients further constrain our ability to draw robust conclusions.

This study highlights the importance of comprehensive stroke management strategies that consider not only clinical factors but also socioeconomic and logistical considerations. Further research addressing the identified limitations is warranted to inform policy and practice aimed at improving stroke care delivery in our region.

## Conclusion

This research, the first of its kind in a Latin American setting, evaluated the amount of DALYs among AIS patients treated with MT. A substantial gap in disability-free life years between patients referred and non-referred patients was detected, with the former experiencing an average additional loss of 9.51 years. Despite initial socioeconomic and demographic differences, adjustments for confounding factors made the outcomes comparable, emphasizing the importance of prioritizing interhospital referral strategies for acute stroke treatment, particularly with timely referral times.

Additionally, it offered insights into the associated cost-effectiveness ratio using real billing data, marking an essential step in understanding the efficacy, safety, and feasibility of secondary MT referral in our context. Furthermore, our sociodemographic analysis sheds light on factors perpetuating the asymmetry in stroke incidence and burden across different socioeconomic groups.

Lastly, this study lays the groundwork for identifying obstacles and strengths for MT referral and sets the stage for future local studies aimed at better understanding stroke burden, demographic profiles, treatment strategies and responses to MT. These efforts will inform economic modeling studies and advocate for optimized stroke management practices, contributing to improved care delivery and patient outcomes.

## Data Availability

Data supporting this study is not publicly available due to its clinical history nature and was exclusively used under strict anonymization with authorization of the Ethics Committee at FSFB for this research. For further information contact

## Ethics statement

This research was approved by the Comité Corporativo de Ética de la Investigación at Fundación Santa Fe de Bogotá, on the 24th of July 2023 with the approval number: CCEI-15558-2023.

---

**Appendix 1.** Clinical Characteristics.

| Category | Description | Total (n (%)) | Referred (n) | Non-referred (n) | P-value |
| --- | --- | --- | --- | --- | --- |
| Previous mRS |  |  |  |  |  |
|  | 0 | 32 | 21 (80.8%) | 11 (45.8%) |  |
|  | 1 | 11 | 3 (11.5%) | 8 (33.3%) |  |
|  | 2 | 4 | 1 (3.8%) | 3 (12.5%) |  |
|  | 3 | 3 | 1 (3.8%) | 2 (8.3%) |  |
|  | total | 50 (98%) | 26 | 24 | 0.015''' |
|  | Missing | 1 (2%) |  |  |  |
| mRS upon release |  |  |  |  |  |
|  | 0 | 4 | 2 (8.7%) | 2 (9.1%) |  |
|  | 1 | 5 | 2 (8.7%) | 3 (13.6%) |  |
|  | 2 | 4 | 2 (8.7%) | 2 (9.1%) |  |
|  | 3 | 3 | 3 (13%) | 0 (0%) |  |
|  | 4 | 8 | 4 (17.4%) | 4 (18.2%) |  |
|  | 5 | 5 | 5 (21.7%) | 0 (0%) |  |
|  | 6 | 16 | 5 (21.7%) | 11 (50%) |  |
|  | total | 45 (88.2%) | 23 | 22 | 0.193''''' |
|  | Missing | 6 (11.8%) |  |  |  |
| mRS at follow-up |  |  |  |  |  |
|  | 0 | 0 | 0 (0%) | 0 (0%) |  |
|  | 1 | 8 | 5 (18.5%) | 3 (12.5%) |  |
|  | 2 | 5 | 3 (11.1%) | 2 (8.3%) |  |
|  | 3 | 6 | 4 (14.8%) | 2 (8.3%) |  |
|  | 4 | 4 | 4 (14.8%) | 0 (0%) |  |
|  | 5 | 3 | 1 (3.7%) | 2 (8.3%) |  |
|  | 6 | 25 | 10 (37%) | 15 (62.5%) |  |
|  | total | 51 (100%) | 27 | 24 | 0.055''''' |
|  |  |  | 17.81* (SD 6.058) | 15.21* (SD 5.413) | 0.113' |
|  | total | 51 (100%) | 27 | 24 |  |
| NIHSS score upon release |  |  |  |  |  |
|  |  |  | 13.23* (SD 9.406) | 5.08* (SD 3.593) | 0.005' |
|  | total | 35 (68.6%) | 22 | 13 |  |
|  | Missing | 16 (31.3%) |  |  |  |
| ASPECTS |  |  |  |  |  |
|  |  |  | 6.95* (SD 1.46) | 8.15* (SD 1.86) |  |
|  | total | 33 (64.7%) | 20 | 13 | 0.044''''' |
|  | Missing | 18 (35.2%) |  |  |  |
| Obstruction site |  |  |  |  |  |
|  | Internal carotid artery | 3 (5.9%) | 3 (11.1%) | 0 (0%) |  |
|  | Left medial cerebral artery M1-M2 | 21 (41.2%) | 10 (37%) | 11 (45.8%) |  |
|  | Left medial cerebral artery M3 | 3 (5.9%) | 2 (7.4%) | 1 (4.2%) |  |
|  | Right medial cerebral artery M1-M2 | 9 (17.6%) | 6 (22.2%) | 3 (12.5%) |  |
|  | Right medial cerebral artery M3 | 2 (3.9%) | 0 (0%) | 2 (8.3%) |  |
|  | Posterior circulation | 8 15.7%) | 4 (14.8%) | 4 (16.7%) |  |
|  | Other | 5 (9.8%) | 2 (7.4%) | 3 (12.5%) |  |
|  | total | 51 (100%) | 27 | 24 | 0.410''''' |
| Prior IVT to MT |  |  |  |  |  |
|  | Yes | 28 (54.9%) | 14 (51.9%) | 14 (58.3%) | 0.642'' |
|  | total | 51 (100%) | 27 | 24 |  |
| TICI score after MT |  | 2.88* (DE 1.25) | 2.88* (DE 1.25) | 2.87* (1.29) |  |
|  | total | 50 (98.4%) | 27 | 23 | 0.969"" |
|  | Missing | 1 (1.96%) |  |  |  |
| Post IVT complications |  |  |  |  |  |
|  | Non-symptomatic intracranial hemorrhage | 1 | 1 (7.1%) | 0 (0%) |  |
|  | Symptomatic intracranial hemorrhage | 2 | 1 (7.1%) | 1 (7.1%) |  |
|  | Major systemic hemorrhage | 0 | 0 | 0 (0%) |  |
|  | Angioedema | 1 | 1 (7.1%) | 0 (0%) |  |
|  | Reperfusion lesion | 0 | 0 | 0 (0%) |  |
|  | Death | 2 | 0 (0%) | 2 (15.4%) |  |
|  | Other | 2 | 1 (7.1%) | 1 (7.1%) |  |
|  | None | 19 | 10 (71.4%) | 9 (69.2%) |  |
|  | total | 27 (96.4%) | 14 | 13 | 0.546" |
|  | Missing | 1 (3.6%) |  |  |  |
| Post MT complications |  |  |  |  |  |
|  | Non-symptomatic intracranial hemorrhage | 2 | 1 (3.7%) | 1 (4.2%) |  |
|  | Symptomatic intracranial hemorrhage | 3 | 1 (3.7%) | 2 (8.3%) |  |
|  | Embolism to other vascular territories | 0 | 0 | 0 |  |
|  | Vascular dissection | 0 | 0 | 0 |  |
|  | Vasospasm | 0 | 0 | 0 |  |
|  | Associated to stent | 0 | 0 | 0 |  |
|  | Death | 0 | 0 | 0 |  |
|  | Other | 0 | 0 | 0 |  |
|  | None | 46 | 25 (92.6%) | 24 (87.5%) |  |
|  | total | 51 (100%) | 27 | 24 | 0.776" |
| Complications during hospital stay |  |  |  |  |  |
|  | Urinary tract infections | 2 | 1 (3.7%) | 1 (4.2%) |  |
|  | Pneumonia / nosocomial infection | 4 | 2 (7.4%) | 2 (8.3%) |  |
|  | Allergic reaction | 0 | 0 (0%) | 0 (0%) |  |
|  | Tracheitis | 0 | 0 (0%) | 0 (0%) |  |
|  | Operatory site infection | 0 | 0 (0%) | 0 (0%) |  |
|  | Death | 15 | 6 (22.2%) | 9 (37.5%) |  |
|  | Decompressive craniotomy requirement | 0 | 0 (0%) | 0 (0%) |  |
|  | Hematoma in puncture site | 0 | 0 (0%) | 0 (0%) |  |
|  | None | 30 | 18 (66.6%) | 12 (50%) |  |
|  | total | 51 (100%) | 27 | 24 | 0.699" |
| Death (on follow-up) |  |  |  |  |  |
|  | Yes | 25 (49%) | 10 (37%) | 15 (62.5%) |  |
|  | total | 51 (100%) | 27 | 24 | 0.069" |
| Cause of death |  |  |  |  |  |
|  | Ischemic stroke | 19 | 10 (100%) | 9 (64.3%) |  |
|  | Pulmonary thromboembolism | 0 | 0 (0%) | 0 (0%) |  |
|  | Respiratory failure | 0 | 0 (0%) | 0 (0%) |  |
|  | Major systemic hemorrhage | 0 | 0 (0%) | 0 (0%) |  |
|  | Symptomatic intracranial hemorrhage | 0 | 0 (0%) | 0 (0%) |  |
|  | Systemic infection | 1 | 0 (0%) | 1 (7.1%) |  |
|  | Another** | 4 | 0 (0%) | 4 (28.6%) |  |
|  | total | 24 (47.1%) | 10 | 14 | 0.105" |
|  | Missing | 11 (52.9%) |  |  |  |

\* = average. ST = standard deviation. ' = T- test (two-tailed). " = "Chi-squared Pearson asymptotic (two-tailed). "' = Exact Mann-Whitney test (two-tailed). "" = Exact Pearson chi-squared test (two-tailed). \*\*represent multifactorial causes such as cardiogenic shock, acute coronary syndrome, tracheostomy disfunction, broncho aspiration and acute renal failure.

**Appendix 2.**
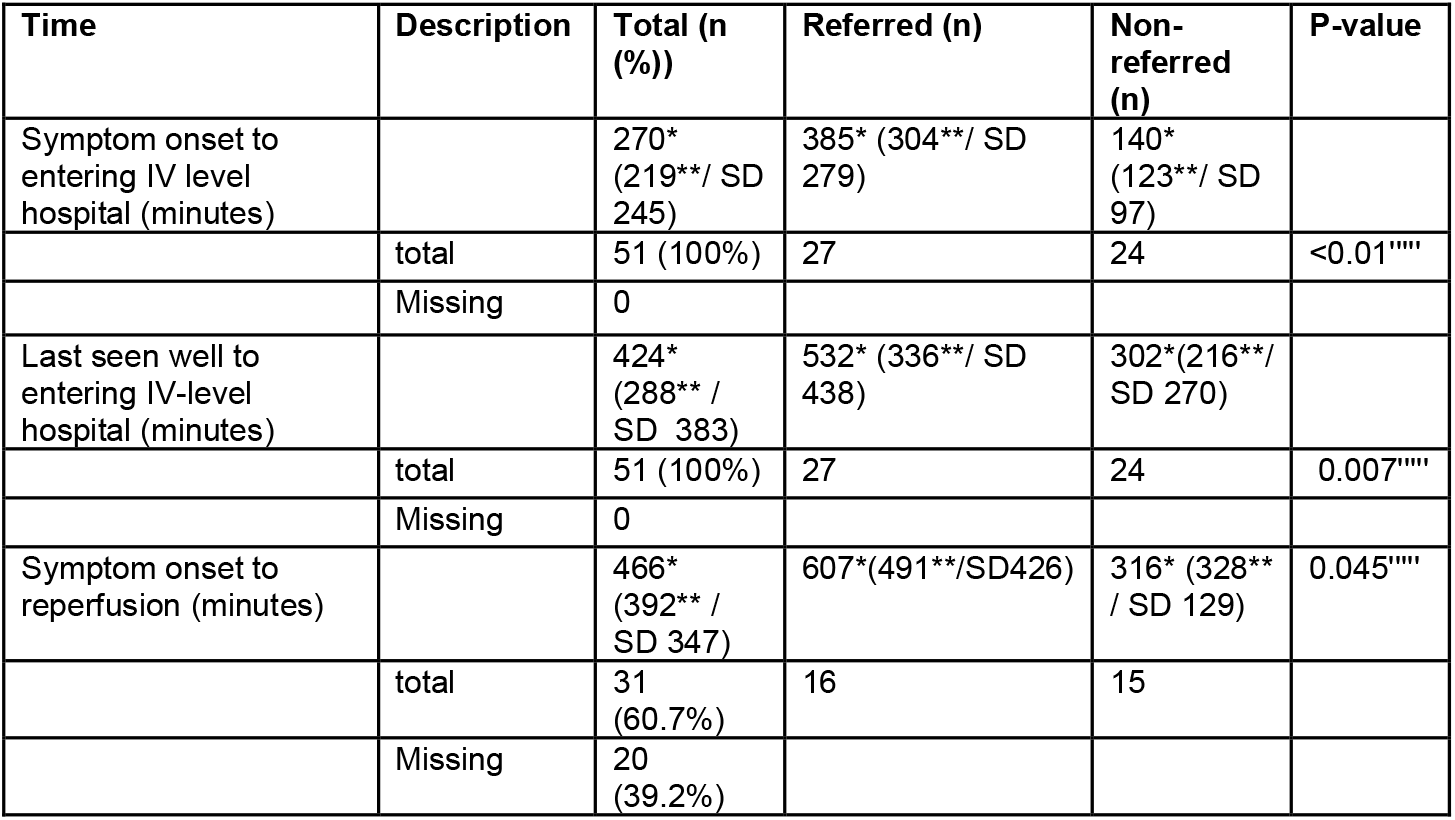
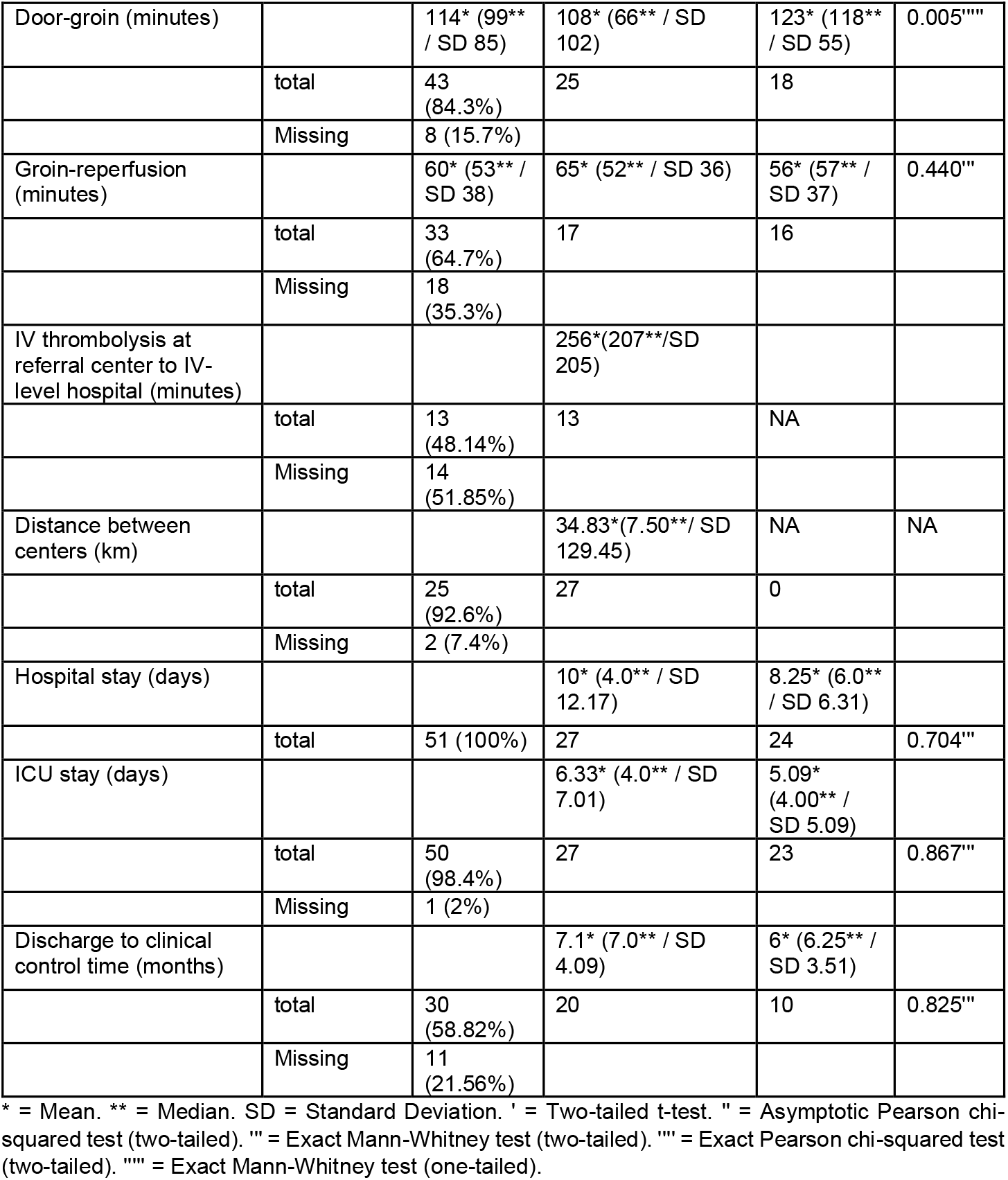
Average intervention times per group.

**Appendix 3.** Ordinal nonparametric regression model (variables without normality) for the difference between the accumulation of DALYs (adjusted and unadjusted model).

| Model | P-value | Estimation (beta) | 95% confidence intervals (CI) |
| --- | --- | --- | --- |
| Difference between the accumulation of DALYs (referred vs non-referred). | 0.003 | 2.433 | (0.813- 4.054) |
| Adjusted difference between the accumulation of DALYs (referred vs non-referred). ** | 0.147 | 7.280 | (-2.558- 17.118) |

\*\*Adjusted for previous baseline mRS, arterial hypertension, DM2, time from onset of symptoms to admission to Level IV Hospital / less than 6 hours.

